# Psychometric Properties of the Short-Form Five Facet Mindfulness Questionnaire (FFMQ-15) in Medical Sciences Students: A Cross-Cultural Validation Study with Diagnostic Utility Analysis

**DOI:** 10.64898/2026.10.14.26365572

**Authors:** Omid Parvizifard, Aliakbar Parvizifard, Aliakbar Forughi, Mohammad Mahdi Heidari

## Abstract

**Objective:** Mindfulness is a robust predictor of mental health, yet the psychometric properties of the short-form Five Facet Mindfulness Questionnaire (FFMQ-15) have not been examined in Iranian populations. This study evaluated the psychometric properties, factor structure, and diagnostic utility of the FFMQ-15 among medical sciences students.

**Participants:** A total of 348 students (178 females, 170 males) from Kermanshah University of Medical Sciences participated.

**Methods:** Participants completed the FFMQ-15, Penn State Worry Questionnaire (PSWQ), Difficulties in Emotion Regulation Scale (DERS), and Emotion Regulation Questionnaire (ERQ). Data were analyzed using exploratory and confirmatory factor analyses, composite reliability, and ROC curve analysis.

**Results:** Both EFA and CFA supported the five-factor model, revealing a culturally unique item loading pattern. The FFMQ-15 demonstrated strong convergent and discriminant validity. ROC analysis indicated acceptable diagnostic accuracy for distinguishing high- and low-worry groups (AUC = 0.72, 95% CI: 0.66–0.78).

**Conclusions:** The FFMQ-15 demonstrated acceptable psychometric properties in Iranian medical students. The non-reactivity facet exhibited cultural variability, suggesting that cross-cultural adaptation is essential. The instrument shows promise for screening in high-stress educational settings.

## Introduction

Over the past two decades, mindfulness has emerged as a foundational construct in clinical psychology, psychiatry, and behavioral medicine. The exponential growth in mindfulness research—from approximately 180 publications in 2010 to over 1,400 in 2022—reflects its increasing recognition as both a therapeutic mechanism and a protective factor for mental health [1, 2]. Mindfulness is operationally defined as “the awareness that emerges through paying attention on purpose, in the present moment, and non-judgmentally to the unfolding of experience moment by moment” [3].

Mindfulness-based interventions, including Mindfulness-Based Stress Reduction (MBSR) [4], Mindfulness-Based Cognitive Therapy (MBCT) [5], Acceptance and Commitment Therapy (ACT) [6], and Dialectical Behavior Therapy (DBT) [7], have demonstrated efficacy across a wide spectrum of psychological disorders, including anxiety, depression, chronic pain, and stress-related conditions [8, 9].

### The Five Facet Mindfulness Questionnaire (FFMQ)

Among available instruments, the Five Facet Mindfulness Questionnaire (FFMQ), developed by Baer et al. [10], has become one of the most widely used measures. It assesses five facets: *observing, describing, acting with awareness, non-judging of inner experience*, and *non-reactivity to inner experience*. The 15-item short form (FFMQ-15), developed by Gu et al. [11], retains the five-factor structure with reduced respondent burden.

### The Cross-Cultural Imperative

Despite its widespread use, the cross-cultural validity of the FFMQ-15 remains an area of active investigation. Culture influences both the experience and expression of mindfulness, particularly facets such as non-reactivity and observing, which may be interpreted differently across individualistic versus collectivistic societies [12, 13]. A systematic review by Karl et al. [13] revealed significant measurement non-invariance across 16 countries, underscoring the necessity of validating the FFMQ-15 in each new cultural context.

### The Unique Context of Medical Sciences Students

Medical sciences students constitute a uniquely high-stress population. Studies have consistently shown that medical students report higher levels of psychological distress, anxiety, and depression compared to age-matched peers [14, 15]. Dyrbye et al. [14] documented that medical students experience burnout and emotional exhaustion at rates significantly higher than the general population. Mindfulness has been identified as a protective factor against burnout and emotional exhaustion in healthcare trainees [16, 17].

### Gaps in the Literature

In Iran, psychometric evaluation of the long-form FFMQ-39 has been conducted by Khanjani et al. [18] in 571 university students, confirming the five-factor structure with acceptable fit indices. However, to date, **no study has examined the psychometric properties of the FFMQ-15 in the Iranian population**—a critical gap that limits both international comparative research and the use of this efficient instrument in Iranian clinical and educational settings.

**Novelty and Contribution of the Present Study**

This study addresses several critical gaps:

1. **First psychometric validation of the FFMQ-15 in Iran**—Extending previous work on the long-form, this study provides the first evaluation of the 15-item version.
2. **Focus on medical sciences students**—A high-priority population for occupational health and educational interventions.
3. **Comprehensive validity evidence**—Beyond confirming the five-factor structure, we examine convergent, discriminant, and diagnostic validity.
4. **Cross-cultural contribution**—We investigate whether the factor structure replicates in a collectivistic context.
5. **Clinical and research implications**—The FFMQ-15 can serve as a brief screening tool in educational and clinical settings.

### Research Aims and Hypotheses

1. **Hypothesis 1:** The five-factor model of the FFMQ-15 demonstrates acceptable fit indices in Iranian medical sciences students.
2. **Hypothesis 2:** The FFMQ-15 demonstrates convergent and discriminant validity through theoretically expected correlations with emotion regulation and worry.
3. **Hypothesis 3:** The FFMQ-15 demonstrates acceptable diagnostic utility in distinguishing between high- and low-worry groups.

## Methods

### Study Design and Population

This cross-sectional validation study included all students enrolled at Kermanshah University of Medical Sciences during the 2023–2024 academic year.

### Sample and Sampling Method

A total of 348 students (178 females, 170 males) were recruited through convenience sampling from medicine, paramedical sciences, pharmacy, and dentistry. Sample size exceeded the minimum of 10 participants per item for factor analysis [19]. Following ethics committee approval (IR.KUMS.MED.REC.1401.054) and obtaining written informed consent, questionnaires were administered in classroom settings.

### Research Instruments

1. **Five Facet Mindfulness Questionnaire-Short Form (FFMQ-15)**—A 15-item instrument assessing five facets of mindfulness on a 5-point Likert scale [10, 11]. Items for acting with awareness and non-judging are reverse-scored. Persian translation was performed using the forward-backward translation method [20].
2. **Penn State Worry Questionnaire (PSWQ)**—A 16-item instrument assessing the tendency to worry. The Persian version has demonstrated reliability (Cronbach’s alpha = 0.86) [21].
3. **Difficulties in Emotion Regulation Scale (DERS)**—A 36-item instrument measuring six dimensions of emotion regulation difficulties. The Persian version has confirmed psychometric properties [22].
4. **Emotion Regulation Questionnaire (ERQ)**—A 10-item instrument assessing cognitive reappraisal and expressive suppression [23]. Persian validation has reported Cronbach’s alpha coefficients ranging from 0.81 to 0.91 [24].

### Data Analysis

Data were analyzed using SPSS version 25, LISREL 8.8, and MedCalc. The analytical strategy included:

1. **Exploratory Factor Analysis (EFA)**—Principal component analysis with varimax rotation to explore the factor structure in this cultural context.
2. **Confirmatory Factor Analysis (CFA)**—Maximum likelihood estimation to evaluate model fit using multiple indices (χ^2^/df, GFI, AGFI, CFI, IFI, RMSEA). Acceptable fit was defined as: CFI ≥ 0.80, GFI ≥ 0.90, AGFI ≥ 0.85, RMSEA ≤ 0.08 [25-27].
3. **Reliability**—Cronbach’s alpha and composite reliability (CR).
4. **Convergent and Discriminant Validity**—Pearson correlations between FFMQ-15 and criterion measures.
5. **Diagnostic Utility**—ROC curve analysis to determine the optimal cutoff score for distinguishing between high- and low-worry groups (based on PSWQ scores > 65).

## Results

### Demographic Characteristics

Of the 348 participants, 51.1% (178) were female and 48.9% (170) were male. Representation across disciplines: medicine (44.5%, 155), paramedical sciences (23.6%, 82), pharmacy (17.8%, 62), and dentistry (14.1%, 49). The mean age was 22.4 years (SD = 3.1, range = 18–35).

### Exploratory Factor Analysis

The Kaiser-Meyer-Olkin (KMO) measure was 0.79, and Bartlett’s test of sphericity was significant (χ^2^ = 1083.77, df = 105, p < 0.001), confirming sampling adequacy. EFA extracted five factors with eigenvalues > 1, accounting for 58.4% of the total variance. Notably, items 5, 10, and 15 (non-reactivity facet) loaded on the same factor but demonstrated lower loadings (0.32–0.45) compared to other facets, suggesting cultural variability in the interpretation of this construct. Items for acting with awareness (3, 8, 13) and non-judging (4, 9, 14) consistently loaded on their respective factors, supporting the robustness of these facets across cultures.

### Confirmatory Factor Analysis

The five-factor model was tested using CFA. Fit indices are presented in Table 1. The GFI value of 0.92 indicated good fit, RMSEA = 0.073 was within the acceptable range, and CFI = 0.82, while below the conventional 0.90 threshold, is considered acceptable in psychological research with complex models [26, 27]. The χ^2^/df ratio (13.3) was attributed to the sensitivity of the chi-square statistic to sample size [19].

**Table 1.** Fit Indices for the Five-Factor Model of the FFMQ-15.

| Index | Value | Index | Value |
| --- | --- | --- | --- |
| $\chi^2$ | 82.250 | **GFI** | 0.92 |
| **df** | 13 | **AGFI** | 0.88 |
| **P-value** | 0.000 | **CFI** | 0.82 |
| $\chi^2/\text{df}$ | 13.3 | **RMSEA** | 0.073 |

### Standardized Factor Loadings and Reliability Indices

Standardized factor loadings ranged from 0.32 to 0.84. As shown in Figure 1, the strongest loadings were observed for the describing facet (items 2, 7, 12: λ = 0.72–0.84), while the non-reactivity facet yielded the lowest loadings (items 5, 10, 15: λ = 0.32–0.45), consistent with previous Iranian [18] and international [11] findings **(Table 2)**.

**Table 2.** Standardized Factor Loadings, Composite Reliability, and AVE for FFMQ-15 Facets.

| Facet | Items | Factor Loadings ($\lambda$) | CR | AVE |
| --- | --- | --- | --- | --- |
| Observing | 1, 6, 11 | 0.51–0.62 | 0.62 | 0.35 |
| Describing | 2, 7, 12 | 0.72–0.84 | 0.76 | 0.52 |
| Acting with Awareness | 3, 8, 13 | 0.48–0.61 | 0.58 | 0.31 |
| Non-Judging | 4, 9, 14 | 0.55–0.68 | 0.67 | 0.40 |
| Non-Reactivity | 5, 10, 15 | 0.32–0.45 | 0.52 | 0.26 |

*Note: CR = Composite Reliability; AVE = Average Variance Extracted*

**Figure 1.**
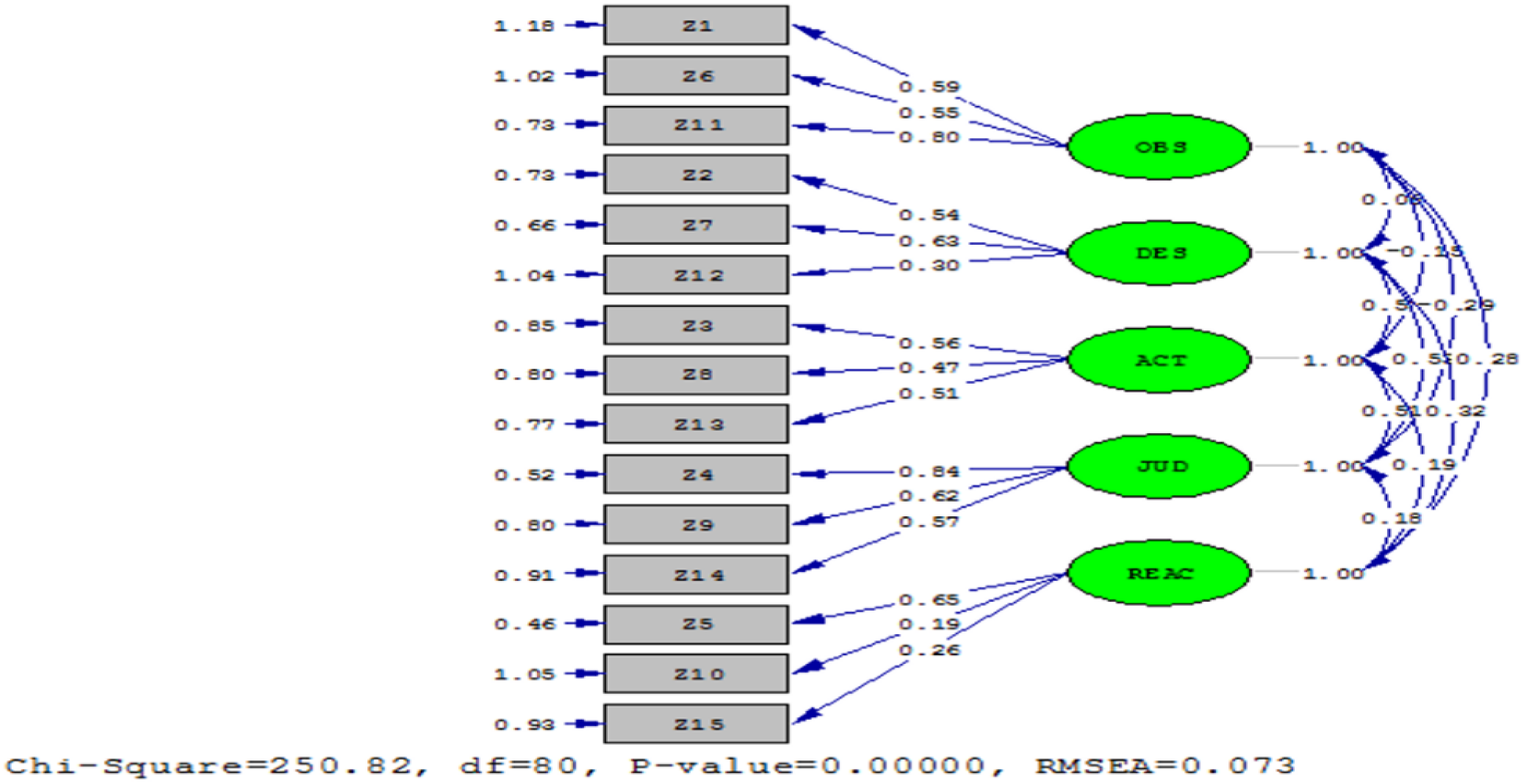
Standardized path diagram of the five-factor measurement model for the FFMQ-15. Note: Rectangles represent observed items (1–15), ovals represent latent factors (Obs = Observing, Des = Describing, Act = Acting with Awareness, NJ = Non-Judging, NR = Non-Reactivity), and circles represent error terms (e1–e15). Numbers on arrows are standardized factor loadings (λ). All factor loadings are significant at p < 0.01. Inter-factor correlations are shown with double-headed arrows.

### Convergent and Discriminant Validity

Table 3 presents the correlations between FFMQ-15 facets and criterion measures at the facet level, providing a more nuanced understanding of the relationship between mindfulness components and emotion regulation.

**Table 3.** Correlations Between FFMQ-15 Facets and Criterion Measures.

| FFMQ-15 Facet | DERS (ER Difficulties) | ERQ-Reappraisal | ERQ-Suppression | PSWQ (Worry) |
| --- | --- | --- | --- | --- |
| Observing | -0.28** | 0.14* | -0.09 | -0.22** |
| Describing | -0.51** | 0.21** | -0.34** | -0.38** |
| Acting with Awareness | -0.48** | 0.08 | -0.19** | -0.41** |
| Non-Judging | -0.44** | 0.15* | -0.22** | -0.35** |
| Non-Reactivity | -0.31** | 0.09 | -0.15* | -0.29** |
| **Total FFMQ-15** | **-0.59\*\*** | **0.17\*\*** | **-0.27\*\*** | **-0.49\*\*** |

*Note: \* $p < 0.05$ , \*\* $p < 0.01$*

This facet-level analysis reveals that the describing facet shows the strongest associations with emotion regulation variables, while the non-reactivity facet demonstrates the weakest relationships, consistent with the lower reliability of this subscale.

### Diagnostic Utility and ROC Curve Analysis

To evaluate the clinical utility of the FFMQ-15, we conducted a receiver operating characteristic (ROC) curve analysis to determine the instrument’s ability to distinguish between students with high versus low levels of worry, based on PSWQ scores (> 65 indicating high worry). The analysis yielded an area under the curve (AUC) of 0.72 (95% CI: 0.66–0.78), indicating moderate diagnostic accuracy. The optimal cutoff score for the FFMQ-15 total score was determined to be 52, which provided a sensitivity of 0.63 and a specificity of 0.69. This indicates that at this threshold, the instrument correctly identified 63% of students with high worry and correctly identified 69% of students without high worry. These findings suggest that the FFMQ-15 holds promise as a brief screening tool for identifying medical sciences students who may be at risk for clinically significant worry and could benefit from further psychological assessment or mindfulness-based interventions.

### Reliability

Cronbach’s alpha for the total scale was 0.63, within the acceptable range for research purposes [28]. Subscale alphas were: observing = 0.58, describing = 0.74, acting with awareness = 0.61, non-judging = 0.69, and non-reactivity = 0.52. The low reliability of the non-reactivity subscale is consistent with findings by Gu et al. [11] (α = 0.54) and is addressed in the discussion.

## Discussion

### Summary of Findings

This study provides the first comprehensive psychometric evaluation of the FFMQ-15 in an Iranian population, specifically among medical sciences students. Overall, the findings indicate that the FFMQ-15 possesses acceptable psychometric properties, supporting its use in non-clinical educational settings. Beyond confirming the five-factor structure (Figure 1), our study uniquely contributes to the literature by (a) conducting both EFA and CFA to examine culture-specific patterns, (b) presenting facet-level correlations with emotion regulation, and (c) evaluating the diagnostic utility of the instrument through ROC curve analysis.

### Factor Structure: Cross-Cultural Insights

The EFA and CFA results (Figure 1) reveal a noteworthy finding: while the five-factor structure of the FFMQ-15 replicates in the Iranian context, the pattern and strength of factor loadings exhibit culture-specific nuances. The non-reactivity facet items (5, 10, 15) consistently showed the lowest loadings (0.32–0.45), a pattern that has been observed in other non-Western populations [12, 13, 29]. This suggests that the construct of “non-reactivity to inner experience” may be less salient or more difficult to conceptualize in collectivistic cultures like Iran.

Christopher et al. [12] found that Thai participants demonstrated different patterns of mindfulness endorsement compared to U.S. participants, emphasizing the role of cultural scripts in reporting mindful experiences. Similarly, Karl et al. [13] documented significant measurement non-invariance across 16 countries, with non-Western samples often exhibiting lower fit indices. Our findings align with these cross-cultural studies, suggesting that mindfulness assessment tools require careful cultural adaptation.

### Comparison with Iranian Studies on FFMQ-39

Our results align closely with previous validation of the long-form FFMQ-39 in Iran. Khanjani et al. [18] reported a CFI of 0.81 and RMSEA of 0.06 in 571 university students, and Tamannaeifar et al. [30] found similar fit indices in Tehran University students. This consistency across versions suggests that the FFMQ-15 retains the essential factor structure of the longer version, supporting its utility as a more efficient alternative.

### Convergent and Discriminant Validity

The strongest aspect of this validation lies in its comprehensive validity evidence. The facet-level correlations (Table 3) reveal a nuanced pattern: the describing and acting with awareness facets demonstrate the strongest associations with emotion regulation difficulties, while the non-reactivity facet shows weaker relationships. This pattern suggests that different facets of mindfulness may contribute differentially to psychological outcomes, which has important implications for intervention design.

The strong negative correlation between FFMQ-15 and DERS (r = -0.59) and PSWQ (r = -0.49) aligns with the extensive literature linking mindfulness to reduced emotional distress and cognitive rigidity [31, 32]. The positive correlation with cognitive reappraisal (r = 0.17) and negative correlation with expressive suppression (r = -0.27) further confirm that mindfulness functions as an adaptive emotion regulation strategy [33].

### Diagnostic Utility: A Novel Contribution

A unique contribution of this study is the evaluation of the diagnostic utility of the FFMQ-15 through ROC curve analysis. The AUC of 0.72 (95% CI: 0.66–0.78) indicates moderate diagnostic accuracy, suggesting that the FFMQ-15 can serve as a useful screening tool in educational settings. The optimal cutoff score of 52 provides a practical threshold for identifying students who may benefit from mindfulness-based interventions.

This finding addresses a gap in the literature, as most validation studies focus solely on construct validity without examining clinical utility. For medical educators and mental health practitioners, this diagnostic evidence strengthens the case for incorporating the FFMQ-15 into routine screening protocols.

### Reliability and Its Interpretation

The internal consistency of the total scale (α = 0.63) is modest but acceptable for brief measures with limited items [28]. This is consistent with findings from other short-form mindfulness measures; for example, Bohlmeijer et al. [34] reported alpha values ranging from 0.60 to 0.78 across subscales. Gu et al. [11] noted that reduced item numbers inherently limit reliability coefficients. Importantly, the robust convergent and discriminant validity evidence substantially mitigates concerns about the modest alpha coefficient, as validity evidence is ultimately more critical than reliability alone [35].

### Clinical and Research Implications

The availability of a validated brief mindfulness measure has practical benefits for:

1. **Screening in educational settings**—The FFMQ-15 can identify students at risk for psychological distress, enabling early intervention.
2. **Intervention evaluation**—Repeated assessments in mindfulness-based interventions, minimizing respondent fatigue.
3. **Cross-cultural research**—Facilitating international comparisons of mindfulness across different national contexts.
4. **Preventive programs**—Supporting university counseling centers in identifying students who may benefit from stress management programs.

### Limitations and Future Directions

This study has several limitations:

1. **Convenience sampling**—Limits generalizability, though the sample’s heterogeneity across four disciplines partially mitigates this concern.
2. **Internal consistency only**—Test-retest reliability was not examined and should be addressed in future research.
3. **Non-clinical sample**—Validation in clinical populations is needed to establish utility in mental health settings.
4. **Non-reactivity facet reliability**—The low α (0.52) suggests that item revision or cultural adaptation may be beneficial.

### Future Research Directions

Future research should consider:

1. Conducting qualitative interviews to understand how Iranian respondents interpret and respond to each item, particularly those assessing non-reactivity.
2. Evaluating test-retest reliability and sensitivity to change following mindfulness-based interventions.
3. Examining measurement invariance across different Iranian demographic groups (gender, age, educational level).
4. Testing competing factor structures (e.g., bifactor models) to determine whether the five-factor structure is optimal for Iranian populations.
5. Comparing FFMQ-15 scores between clinical and non-clinical samples to further establish diagnostic utility.

## Conclusions

The Persian short-form Five Facet Mindfulness Questionnaire (FFMQ-15) demonstrates acceptable validity and modest internal consistency in Iranian medical sciences students. This first validation of the FFMQ-15 in Iran fills a critical gap in the cross-cultural mindfulness literature. The FFMQ-15 represents a practical, efficient tool for mindfulness assessment in research and screening contexts. However, the lower reliability of the non-reactivity facet underscores the need for cultural adaptation and cautious interpretation. Despite these limitations, the FFMQ-15 can be recommended as a cost-effective measure for use in non-clinical educational and research settings in Iran.

## Data Availability

The dataset generated and analyzed during the current study is available from the corresponding author upon reasonable request.

## Declarations

### 1. Ethics Approval and Consent to Participate

This study was conducted **in accordance with the Declaration of Helsinki**. Ethical approval was obtained from the Ethics Committee of Kermanshah University of Medical Sciences (IR.KUMS.MED.REC.1401.054). Written informed consent was obtained from all participants.

### 2. Consent for Publication

Not applicable.

### 3. Availability of Data and Materials

Available from the corresponding author upon reasonable request.

### 6. Authors’ Contributions

OP and AP conceptualized and designed the study. AF and MMH collected and analyzed the data. OP and AP drafted and critically revised the manuscript. All authors approved the final manuscript.

## 7. Acknowledgements

The authors thank all students who participated and the administration of Kermanshah University of Medical Sciences for facilitating data collection.

## References

1. Kabat-Zinn J. Mindfulness-based interventions in context: past, present, and future. Clin Psychol Sci Pract. 2003;10(2):144–56.

2. Goldberg SB, Tucker RP, Greene PA, Davidson RJ, Wampold BE, Kearney DJ, et al. Mindfulness-based interventions for psychiatric disorders: A systematic review and meta-analysis. Clin Psychol Rev. 2018;59:52–60.

3. Kabat-Zinn J. Wherever you go, there you are: Mindfulness meditation in everyday life. New York: Hyperion; 1994.

4. Kabat-Zinn J. Full catastrophe living: Using the wisdom of your body and mind to face stress, pain, and illness. New York: Delta; 1990.

5. Segal ZV, Williams JMG, Teasdale JD. Mindfulness-based cognitive therapy for depression. New York: Guilford Press; 2002.

6. Hayes SC, Strosahl KD, Wilson KG. Acceptance and commitment therapy. New York: Guilford Press; 1999.

7. Linehan MM. Cognitive-behavioral treatment of borderline personality disorder. New York: Guilford Press; 1993.

8. Hofmann SG, Sawyer AT, Witt AA, Oh D. The effect of mindfulness-based therapy on anxiety and depression: A meta-analytic review. J Consult Clin Psychol. 2010;78(2):169–83.

9. Khoury B, Lecomte T, Fortin G, Masse M, Therien P, Bouchard V, et al. Mindfulness-based therapy: A comprehensive meta-analysis. Clin Psychol Rev. 2013;33(6):763–71.

10. Baer RA, Smith GT, Hopkins J, Krietemeyer J, Toney L. Using self-report assessment methods to explore facets of mindfulness. Assessment. 2006;13(1):27–45.

11. Gu J, Strauss C, Crane C, Barnhofer T, Karl A, Cavanagh K, et al. Examining the factor structure of the 39-item and 15-item versions of the Five Facet Mindfulness Questionnaire before and after mindfulness-based cognitive therapy for people with recurrent depression. Psychol Assess. 2016;28(7):791–802.

12. Christopher MS, Charoensuk S, Gilbert BD, Neary TJ, Pearce KL. Mindfulness in Thailand and the United States: A case of apples versus oranges? J Clin Psychol. 2009;65(6):590–612.

13. Karl JA, Fischer R, Jose PE. The Five Facet Mindfulness Questionnaire across cultures: A systematic review and meta-analysis of measurement invariance. Eur J Psychol Assess. 2020;36(3):499–510.

14. Dyrbye LN, Thomas MR, Shanafelt TD. Systematic review of depression, anxiety, and other indicators of psychological distress among U.S. and Canadian medical students. Acad Med. 2006;81(4):354–73.

15. Rotenstein LS, Ramos MA, Torre M, et al. Prevalence of depression, depressive symptoms, and suicidal ideation among medical students: A systematic review and meta-analysis. JAMA. 2016;316(21):2214–36.

16. Shapiro SL, Schwartz GE, Bonner G. Effects of mindfulness-based stress reduction on medical and premedical students. J Behav Med. 1998;21(6):581–99.

17. Greeson JM, Toohey MJ, Pearce MJ. An adapted, four-week mind-body skills group for medical students: A pilot study. Med Teach. 2015;37(3):283–90.

18. Khanjani S, Foroughi AA, Bazani M, Rafiee S, Tamannaeifar SH, Habibi M. Psychometric properties of Persian version of five facets of mindfulness questionnaire. J Res Med Sci. 2022;27:29.

19. Kline RB. Principles and practice of structural equation modeling. 4th ed. New York: Guilford Press; 2016.

20. Brislin RW. Back-translation for cross-cultural research. J Cross-Cult Psychol. 1970;1(3):185–216.

21. Shirinzadeh Dastgiri S. Comparison of meta-cognitive and responsibility beliefs between patients with obsessive-compulsive disorder, generalized anxiety disorder, and normal population. Iran J Psychiatry Clin Psychol. 2008;14(1):46–55. [Persian]

22. Edrissi F, Khodadadi M, Bahrainian SA. Structural model of emotion regulation and GAD symptoms. Clin Psychol Stud. 2014;4(15):203–26. [Persian]

23. Gross JJ, John OP. Individual differences in two emotion regulation processes: Implications for affect, relationships, and well-being. J Pers Soc Psychol. 2003;85(2):348–62.

24. Ghasemi Jobaneh R, Arabzadeh F, Jalili Nikoo S, Mohammadi Pour M, Mohsenzadeh F. Validity and reliability of the Persian version of the short form of Freiburg Mindfulness Inventory. J Rafsanjan Univ Med Sci. 2015;14(2):137-50. [Persian]

25. Hu LT, Bentler PM. Cutoff criteria for fit indexes in covariance structure analysis. Struct Equ Modeling. 1999;6(1):1–55.

26. Marsh HW, Hau KT, Wen Z. In search of golden rules: Comment on hypothesis-testing approaches to setting cutoff values for fit indexes. Struct Equ Modeling. 2004;11(3):320–41.

27. Marsh HW, Balla JR, McDonald RP. Goodness-of-fit indexes in confirmatory factor analysis: The effect of sample size. Psychol Bull. 1988;103(3):391–410.

28. DeVellis RF. Scale development: Theory and applications. 4th ed. Thousand Oaks: SAGE Publications; 2017.

29. Wang Y, Xu W, Luo F. Psychometric properties of the Five Facet Mindfulness Questionnaire-20 in Chinese nursing students. Front Psychol. 2021;12:678–89.

30. Tamannaeifar S, Asgharnejad Farid A, Mirzaei M, Soleimani M. Psychometric properties of the Five Facet Mindfulness Questionnaire. Dev Psychol (Iran Psychol). 2019;12(47):321-9. [Persian]

31. Brown KW, Ryan RM. The benefits of being present: Mindfulness and its role in psychological well-being. J Pers Soc Psychol. 2003;84(4):822–48.

32. Shapiro SL, Carlson LE, Astin JA, Freedman B. Mechanisms of mindfulness. J Clin Psychol. 2006;62(3):373–86.

33. Hayes AM, Feldman G. Clarifying the construct of mindfulness in the context of emotion regulation and the process of change in therapy. Clin Psychol Sci Pract. 2004;11(3):255–62.

34. Bohlmeijer E, ten Klooster PM, Fledderus M, Veehof M, Baer R. Psychometric properties of the five facet mindfulness questionnaire in depressed adults and development of a short form. Assessment. 2011;18(3):308–20.

35. Boateng GO, Neilands TB, Frongillo EA, Melgar-Quiñonez HR, Young SL. Best practices for developing and validating scales for health, social, and behavioral research: A primer. Front Public Health. 2018;6:149.

